# Effect of a multi-faceted Rapid Response System re-design on repeat calling of the Rapid Response Team

**DOI:** 10.1101/2020.11.24.20238238

**Authors:** Richard Chalwin, Amy Salter, Jonathon Karnon, Victoria Eaton, Lynne Giles

**Author notes:** ***Corresponding Author*** Richard Chalwin, School of Public Health, Faculty of Health and Medical Sciences, University of Adelaide, Adelaide, SA 5000, Australia.

## Abstract

**Background:** Repeat Rapid Response Team (RRT) calls are associated with increased mortality risk to patients and pose a resource burden to organisations. Use of Non-Technical Skills (NTS) at calls has the potential to reduce potentially preventable repeat calling. NTS are usually improved through training, however this consumes time and financial resources. Modifications to the Rapid Response System (RRS) that promote use of NTS are worth exploring as a cost-effective alternative.

**Methods:** A pre-post observational study of a RRS re-design on proportion of admissions each month subject to repeat RRT calling and number of calls per admission, with univariate and multivariable interrupted time series analyses comparing outcomes between study phases.

**Results:** The proportion of admissions with repeat calls each month increased across both phases of the study period, but the increase was lower in the post re-design phase (change in adjusted regression slope -0.12 (standard error 0.07) post versus pre re-design). The multivariable model showed an estimated 6.0% reduction (P=0.19) in the proportion of admissions having repeat calls at the end of the study versus that predicted had the re-design not occurred.

For the number of calls per the multivariable model predicted a reduction of 0.07 calls per admission at the end of the post re-design phase (95% confidence interval -0.23 – 0.08, P=0.35), equating to one fewer repeat call per 14 patients having RRT calls.

**Conclusion:** This study showed no observed statistically significant effect on rates of repeat calling or numbers of repeat calls per admission from the implementation of a RRS re-design. However, the results demonstrate, at an organisational level, the feasibility of a low-cost initiative to improve NTS use by the RRS.

## INTRODUCTION

Over the past quarter century, the Rapid Response Team (RRT) has evolved from a conceptual advancement of the response to in-hospital cardiac arrest to become a ubiquitous patient safety mechanism. [1,2] Throughout this time, studies and reviews of Rapid Response System (RRS) activity have consistently demonstrated increasing rates of RRT calling. [2-5]

In some respects, this suggests desirable awareness and utilisation of a patient safety mechanism. Indeed, the increase in RRS usage within an organisation has been associated with improved patient survival statistics. [3,6,7] However, ever expanding RRS activity poses a logistical and resourcing burden for hospitals, as most RRTs tend to not be supernumerary, with staff rostered from other substantive roles. [4,8,9] Although adverse effects have not yet been attributed to team members leaving other duties to attend RRT calls, [10] the potential exists for these to occur. This risk could be magnified during concurrent RRT calls as resources are typically not available to provide a full response to more than one call simultaneously. [9]

In this environment, the RRS should seek efficiencies to optimise RRT availability given the need to promptly attend all unexpected clinical deteriorations. One avenue could be through reduction of potentially preventable repeat calling, that is the RRT attending a patient more than once due to inadequate resolution of an initial call, especially when the repeat call closely follows the first. Previously, the authors found increased mortality risk in patients re-attended by the RRT within 24 hours of a previous completed call in which clinical issues remained unresolved. [11]

Deficits in non-technical skills (NTS), such as communication and cooperation, at RRT calls have been identified as a risk factor for potentially preventable repeat calling. [11-13] Effective employment of NTS are crucial due to the inherent time and clinical pressures imposed by the deteriorating patient. [14,15] Ideally, NTS would be augmented by delivery of specialised, simulated scenario training for RRTs. [14, 16] However, such training requires taking staff away from clinical duties, which is often not feasible in resource-limited hospitals.

Therefore, the authors conducted a comprehensive, multi-faceted RRS re-design, aiming to enhance use of NTS at RRT calls, without the need for dedicated training. The re-design drew on themes from the TeamSTEPPS® program, [17,18] and previous research which described RRS improvement initiatives. [14,19-21] The present study builds on this prior work through the novel application of Interrupted Time Series analysis to investigate the effects of the re-design of an existing RRS.

## METHODS

A pre-post intervention study assessing the proportion of patients subject to repeat calling before and after implementation of a RRS re-design was used. The present study completes reporting of the *Impact of Non-Technical Skills on Performance and Effectiveness of a Medical Emergency Team* project (ClinicalTrials.gov: NCT01551160), previous stages of which have already been published. [11-13]

### Participants

Patients attended by the RRT at a tertiary, outer metropolitan hospital between 1^st^ July 2009 and 30^th^ June 2019 were identified from RRS records. Those who were not admitted to the hospital (e.g. day procedures, outpatients, or visitors) and patients under 18 years of age were ineligible for inclusion.

The cohort of in-patient admissions who were attended by the RRT were divided into two groups: those attended by the RRT more than once during an admission (the Repeat Call group) and those with only one RRT call.

Clinical staff were classified into two groups: those rostered to attend calls as part of the RRT (‘members’), and those who recognise clinical deterioration and call the RRT (‘users’).

### Intervention

The RRS re-design incorporated three components, described in detail previously. [13] These components targeted the key NTS domains of leadership, communication, and co-operation both within the RRT and between RRT members and users.

#### 1. Regular RRT meetings

Short meetings for RRT members, designed to address **Leadership** and **Cooperation** within the team, were scheduled to occur at the beginning of each shift. The primary purpose of these meetings was to pre-emptively establish each team member’s role and initial task at RRT calls. This approach was designed to avoid spending valuable time doing this at a deteriorating patient’s bedside.

#### 2. Team Role Badges

Each member of the RRT was required to wear a badge indicating their role while attending calls. This was designed to reinforce the team **Leadership** role as well as facilitate non-verbal **Communication** of all role designations to RRT members and users present at calls.

#### 3. RRT members-to-users “hand-off” procedure

A structured verbal and written process, aiming to improve **Communication** and **Cooperation** between RRT members and users, was introduced for RRT calls ending with the patient remaining on their ward. This formalised the transfer of primary clinical responsibility from the RRT back to the ward team. In particular, the hand-off process encouraged RRT users to voice any ongoing clinical concerns and have them addressed before the RRT departed.

### Study Phases

There were two phases of data collection, punctuated by the implementation of the RRS re-design as detailed above. Phase 1 comprised five years (2009 – 2014) of retrospective data on RRT operational activity retrieved from the hospital RRS electronic database. These data were collected at commencement of the project and findings have been reported separately. [12] At the end of Phase 2, a further five-years (2014 – 2019) of RRT call data were extracted from RRS records.

Aside from the re-design, the configuration and operations of the RRS did not change over the entire study period (i.e. Phase 1 and 2). In particular, the RRT activation criteria, composition of the RRT and provision of Critical Care services at the investigating hospital remained the same throughout.

### Outcome Measures

RRT call data, obtained from the hospital RRS database, were aggregated at the per-patient-admission level. Variables were then created to indicate if each admission contained repeat calls, or not, and the count of those repeat calls.

The admission-level data were then collapsed by study month, derived from the date of hospital entry, with month 1 representing July 2009, through to month 120 in June 2019. A variable was created to indicate study phase (Phase 1: months 1 – 60, Phase 2: months 61 – 120).

The primary outcome of this study was the proportion of admissions with repeat RRT calls from all admissions with at least one RRT call (per month). This was chosen as an indicator of potentially preventable RRS activity that could be measured throughout both study phases.

The secondary outcome was the mean number of RRT calls per admission (from all admissions with at least one RRT call) to investigate aggregate RRT call load on the hospital.

### Other Variables

Demographic data, captured at time of admission, included age, gender, Indigenous identification and socioeconomic status (expressed as a binary variable for Socio-Economic Indexes for Australia (SEIFA) decile of three or less versus greater than three, derived from the 2016 Postal Area Index of Relative Socio-economic Advantage and Disadvantage) [22]. Hospital admission data included elective vs non-elective admission, Charlson Co-Morbidity Index (CCI) and in-patient length of stay (LOS). Counts of hospital admissions during the study were derived from the hospital activity database.

These variables were similarly aggregated by month to account for variations in hospital activity and casemix over the study period. For each study month, the number of admissions, and the percentage of admissions corresponding to gender, Indigenous identification, SEIFA ≤ 3, and non-elective admissions were derived. The mean age, CCI, and hospital LOS were also calculated for each study month.

### Data Analysis

Monthly hospital activity and aggregated patient demographics were compared between study phases using Mann-Whitney U-tests.

The effect of the re-design was assessed by Interrupted Time Series (ITS) methodology as described by Bernal et al. [23] In general terms, ITS analyses use segmented regression to compare the observed effect of an intervention, introduced at a defined time point, on an outcome to the effect predicted in the absence of the intervention. The ITS technique is particularly useful for studying organisation-level interventions where randomised controlled trials are infeasible. [24,25] Thus, for the pre-post structure of this study, ITS was ideal to quantify the impact of the RRS re-design on the outcomes of interest through the change in coefficients of the fitted regression line at the point of introducing the re-design.

Non-seasonal Auto Regressive Integrated Moving Average (ARIMA) models with a first-order auto-correlation were fit for each outcome variable. Study month was used as the time metric in all models. Initially, simple models were fit that considered only time (study month), phase and the interaction of time and phase (i.e. a different intercept and slope corresponding to the post-intervention phase compared to the pre-intervention phase were allowed for in the regression model – see Figure 2 in Bernal et al [23]). Subsequently, multivariable models that included hospital admission rates, patient demographics and admission characteristics were fit to adjust for any variations between months in hospital activity and casemix over the study phases. The final multivariable model retained variables with a corresponding P-value < 0.1. Sensitivity analyses were undertaken to examine the impact of potential outliers. [26]

**FIGURE 1.**
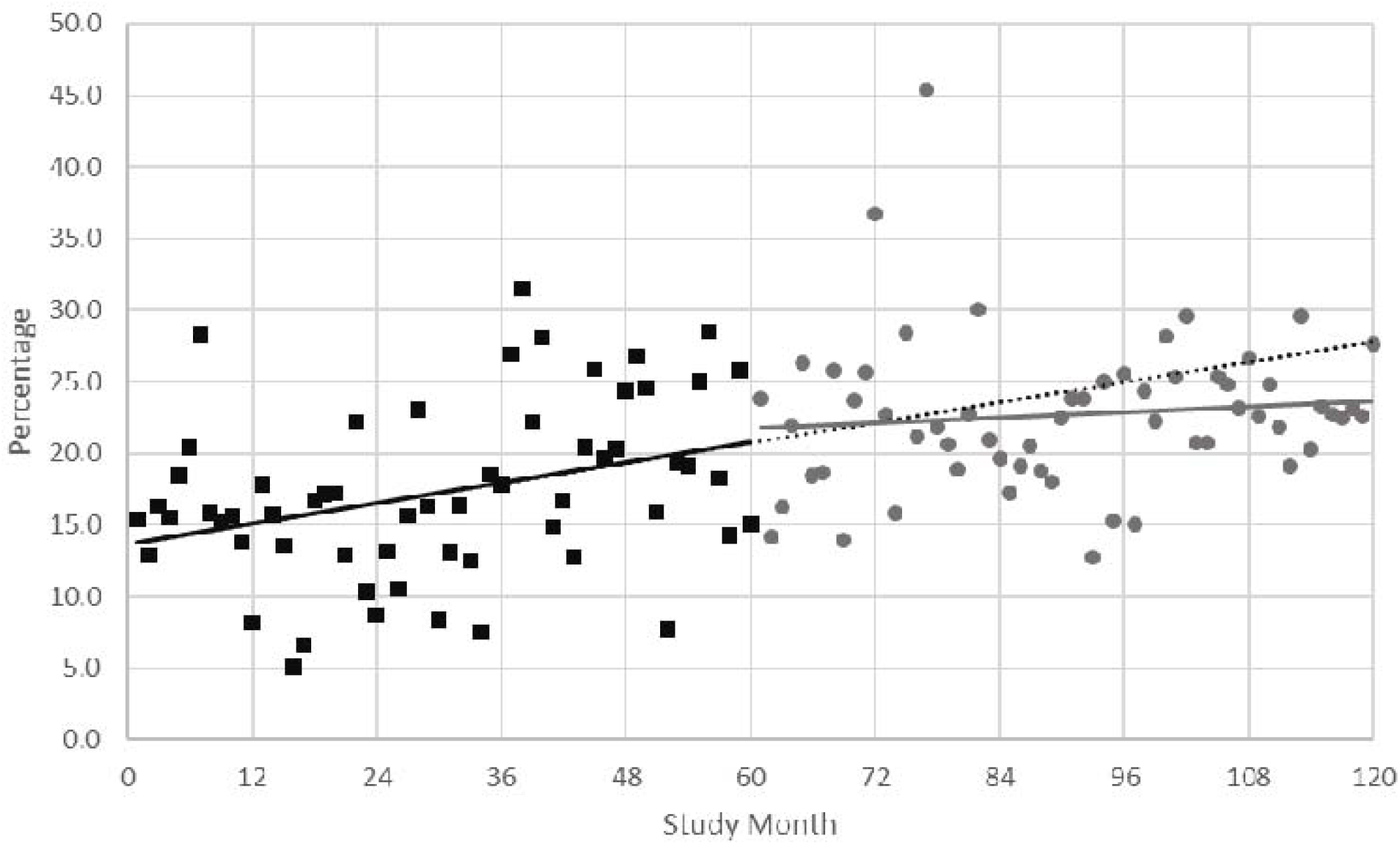
Percentage of repeat call admissions per month representing the ARIMA univariate model. Phase 1 monthly observed data in black squares, with slope illustrated by the solid black line. Phase 2 monthly observed data in grey circles, with slope illustrated by the solid grey line. The slope in Phase 1 is extended into Phase 2 and represented by the dotted black line for comparison with Phase 2 observed data.

**FIGURE 2.**
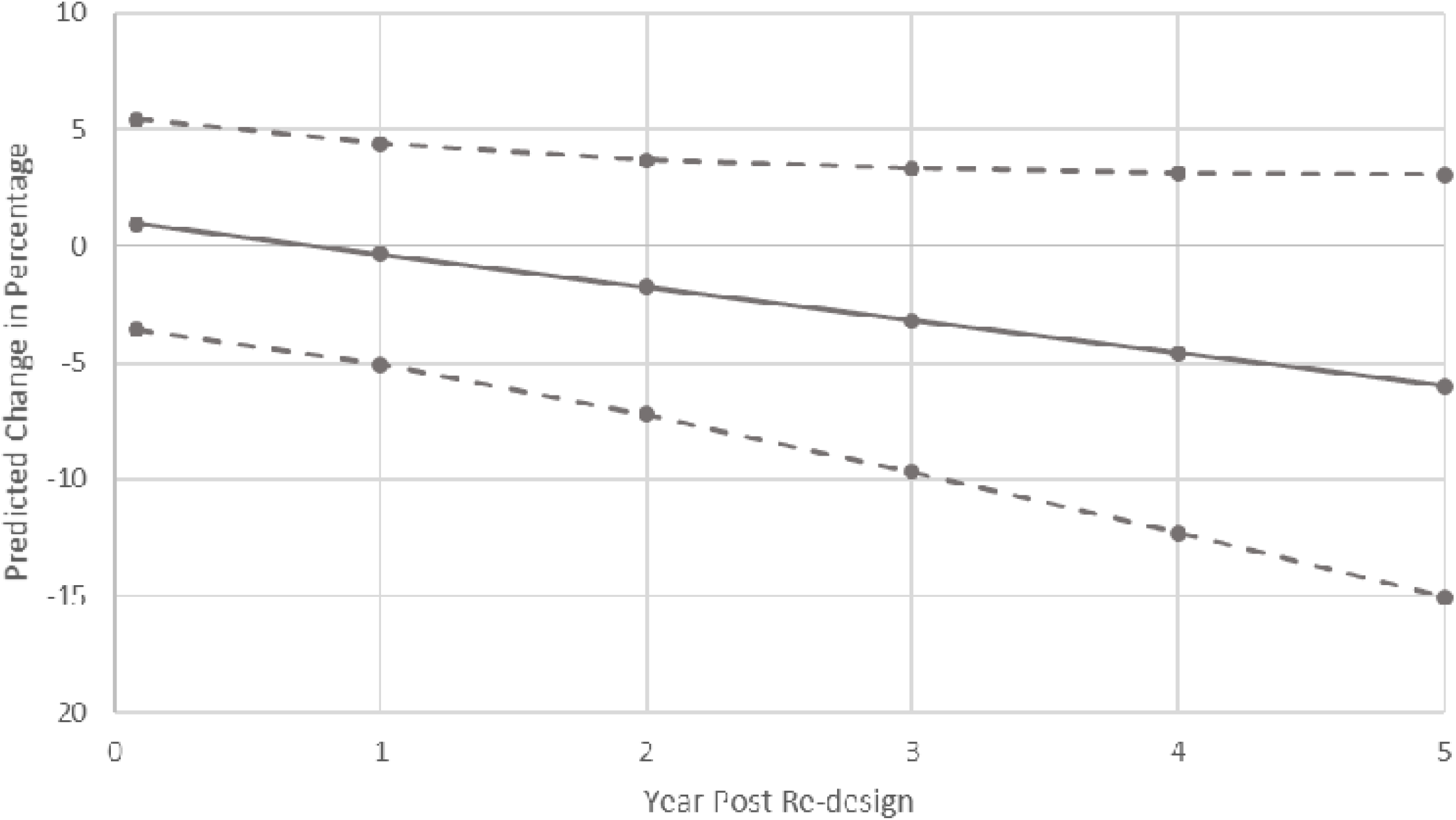
Final Multivariable Model – Cumulative predicted change in percentage of repeat call admissions (per month) associated with the RRS re-design, with 95% confidence intervals.

Predicted changes in the percentage of repeat call admissions and mean number of calls per admission were derived for each year using the approach outlined in the ITS guidance document of the Cochrane Effective Practice and Organisation of Care group. [27] In this way, the cumulative annual changes in the outcome measures that were attributable to the RRS design were estimated. Model fit was assessed by the stationary R^2^ value, where values closer to 1 are indicative of better fit, and the Ljung-Box Q statistic, which indicates if there is a marked lack of fit of the corresponding ARIMA model. [28] Durbin’s alternative statistic was used to assess the extent of auto-correlation in the statistical models. [29]

Statistical analyses were conducted with SPSS (IBM Corp. Released 2019. IBM SPSS Statistics for Windows, Version 26.0. Armonk, NY: IBM Corp), with the exception of Durbin’s alternative statistic, which was calculated using Stata (StataCorp. 2017. Stata Statistical Software: Release 15. College Station, TX: StataCorp LLC).

### Ethics

This study was approved by the Central Adelaide Health Network Human Research Committee (approval number: 2012069).

The need for patient signed consent was waived on the grounds that data used in this study were already collected electronically for hospital quality assurance purposes, no unique patient identifiers were included in the study database, and all individual patient level variables were aggregated by study month prior to analysis and reporting.

## RESULTS

The RRS database provided records for 9754 patients who were attended by the RRT during the study period. From these, 93 paediatric patients and 122 visitors, staff or outpatients were excluded as they were ineligible. A further 12 in-patients for whom the database had incomplete records were also excluded. Of the remaining 9527 patient admissions, 3073 occurred in Phase 1 and 6454 in Phase 2.

Compared to Phase 1, in Phase 2 there were greater mean overall hospital admissions per month (4015 [standard deviation (SD) 419.7] vs 3134 [SD 222.0], P<0.01), shorter mean in-patient LOS (10.9 days [SD 1.6] vs 12.9 [SD 3.0], P<0.01), lower mean patient age (67.4 [SD 2.0] vs 68.6 [SD 2.8], P<0.01), lower percentage of patients with low socioeconomic status (68.2% [SD 7.0%] vs 79.7% [SD 6.4%], P<0.01) and lower mean CCI (4.5 [SD 0.3] vs 4.8 [SD 0.46], P<0.01). Hospital activity and patient demographic data are summarised by year of the study in Table 1.

**Table 1:** Hospital activity and demographic data (for patients having RRT calls) by study year. RRT = Rapid Response Team, SD = standard deviation, LOS = length of stay, SEIFA = socio-economic indexes for Australia, CCI = Charlson co-morbidity index.

| Study Year | Count of All Admissions | Count of RRT Call Admissions | LOS Mean (SD) | Age Mean (SD) | Male Mean % (SD) | Indigenous Mean % (SD) | Low SEIFA Mean % (SD) | CCI Mean (SD) | Non-Elective Mean % (SD) |
| --- | --- | --- | --- | --- | --- | --- | --- | --- | --- |
| 1 | 34238 | 507 | 14.4 (4.2) | 67.8 (3.3) | 48.7 (6.2) | 0.9 (1.3) | 81.7 (6.7) | 4.7 (0.4) | 90.8 (3.1) |
| 2 | 36087 | 506 | 14.0 (3.0) | 69.1 (2.8) | 52.3 (9.4) | 1.8 (2.0) | 79.0 (6.7) | 4.9 (0.5) | 94.4 (2.7) |
| 3 | 37785 | 578 | 11.7 (2.1) | 68.9 (2.6) | 51.4 (7.4) | 3.0 (1.8) | 82.3 (4.5) | 4.8 (0.5) | 93.4 (3.0) |
| 4 | 39441 | 666 | 12.1 (2.2) | 68.0 (3.1) | 50.8 (8.7) | 2.6 (1.7) | 78.4 (8.2) | 4.6 (0.5) | 94.9 (3.2) |
| 5 | 40465 | 816 | 12.6 (2.5) | 69.3 (2.4) | 47.9 (5.2) | 1.7 (1.4) | 77.0 (5.0) | 4.9 (0.4) | 92.9 (2.9) |
| Phase 1 overall | 188016 | 3073 | 12.9 (3.0) | 68.6 (2.8) | 50.2 (7.5) | 2.0 (1.8) | 79.7 (6.4) | 4.8 (0.5) | 93.3 (3.2) |
| 6 | 41098 | 887 | 11.0 (0.6) | 67.2 (2.5) | 52.1 (6.6) | 2.4 (2.1) | 74.5 (5.5) | 4.7 (0.4) | 92.4 (2.7) |
| 7 | 45307 | 1174 | 9.9 (1.5) | 65.8 (2.1) | 45.7 (5.0) | 1.9 (1.2) | 72.4 (6.4) | 4.3 (0.3) | 92.0 (2.8) |
| 8 | 49009 | 1223 | 11.6 (2.0) | 67.8 (1.6) | 49.5 (5.2) | 2.0 (1.6) | 65.5 (5.6) | 4.4 (0.2) | 94.6 (2.6) |
| 9 | 52123 | 1541 | 11.5 (1.8) | 68.2 (1.6) | 49.7 (4.8) | 3.1 (1.7) | 66.6 (4.9) | 4.6 (0.2) | 93.2 (3.3) |
| 10 | 53373 | 1629 | 10.7 (1.3) | 67.9 (1.5) | 49.3 (5.3) | 3.6 (1.8) | 61.8 (4.5) | 4.6 (0.3) | 93.4 (1.9) |
| Phase 2 overall | 240910 | 6454 | 10.9 (1.6) | 67.4 (2.0) | 49.3 (5.6) | 2.6 (1.7) | 68.2 (7.0) | 4.5 (0.3) | 93.1 (2.7) |

### Primary Outcome

The ARIMA univariate model produced a regression coefficient for the slope of 0.115 (standard error (SE) 0.047) in Phase 1, and 0.029 (SE 0.047) in Phase 2, indicating an observed change in slope between phases of -0.087 (SE 0.067).

Similar results were found for the final multivariable model, in which proportion of non-elective admissions and average hospital length of stay were also retained as covariates. In this model, the regression coefficient corresponding to the change in slope due to the re-design was -0.118 (SE 0.067).

The final multivariable model estimated an absolute 6% reduction of RRT attended patients triggering repeat calls (per month) by the fifth year post-implementation (95% confidence interval -15.1 – 3.1, P=0.19), due to the RRS re-design. The estimated cumulative reduction in the observed percentage of repeat call admissions in Phase 2, compared to the percentage predicted if the re-design had not been implemented, is shown in Figure 2.

Durbin’s alternative test statistics were 2.35 on 1 df (P=0.12) and 2.13 on 1 df (P=0.14) for the univariate and multivariable models, respectively, and the stationary R^2^ values were 0.26 for the univariate and 0.30 for the multivariable models, respectively. The Ljung-Box Q statistic indicated there was no significant lack of fit observed for the univariate (15.77 on 17 df; P=0.54), nor for the multivariable model (Q=20.83 on 17 df; P=0.23). Taken together, these statistics suggest the fit of the ITS models is reasonable.

Given the unusual observation in November 2015 (study month 77), a sensitivity analysis was conducted excluding this value. This analysis resulted in slightly attenuated regression coefficients for the univariate (−0.068 [SE 0.064] vs -0.087 [SE 0.067]) and multivariable models (−0.094 [SE 0.065] vs -0.118 [SE 0.067]), and a modest alteration of the absolute reduction in percentage of patients having repeat calls versus predicted to 4.9% (95%CI -13.7 – 3.81, P=0.27) as shown in Supplementary Material 1.

### Secondary Outcome

The change in regression coefficient for the mean number of calls per admission in Phase 2 compared to Phase 1 associated with implementation of the re-design was -0.001 (SE 0.001) in the ARIMA univariate model. Figure 3 shows the observed data for Phase 1 and Phase 2.

**FIGURE 3.**
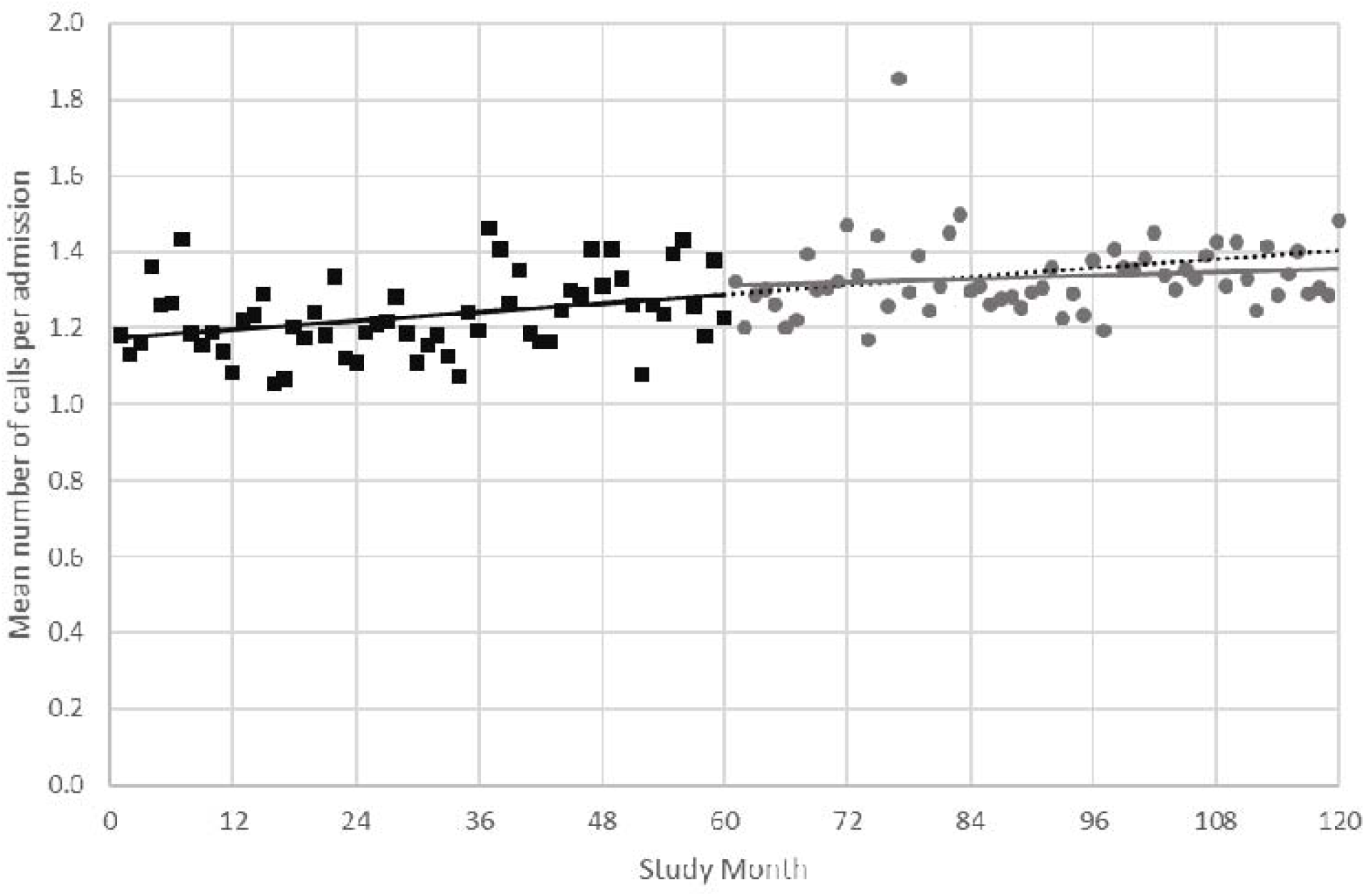
Mean number of calls per admission by study month for the ARIMA univariate model. Phase 1 observed data in black squares, with slope illustrated by the solid black line. Phase 2 observed data in grey circles, with slope illustrated by the solid grey line. The Phase 1 slope is extended into Phase 2 and represented by the dotted black line for comparison with Phase 2 observed data.

At the end of the Phase 2, the final multivariable model, retaining hospital LOS, showed a predicted reduction of 0.07 (95%CI -0.23 – 0.08) calls per admission (p=0.35). Compared to the predicted increase in repeat calls in the absence of the intervention, the results suggest one repeat call would have been avoided for every 14 initial RRT calls in the fifth-year post implementation (see Figure 4).

**FIGURE 4.**
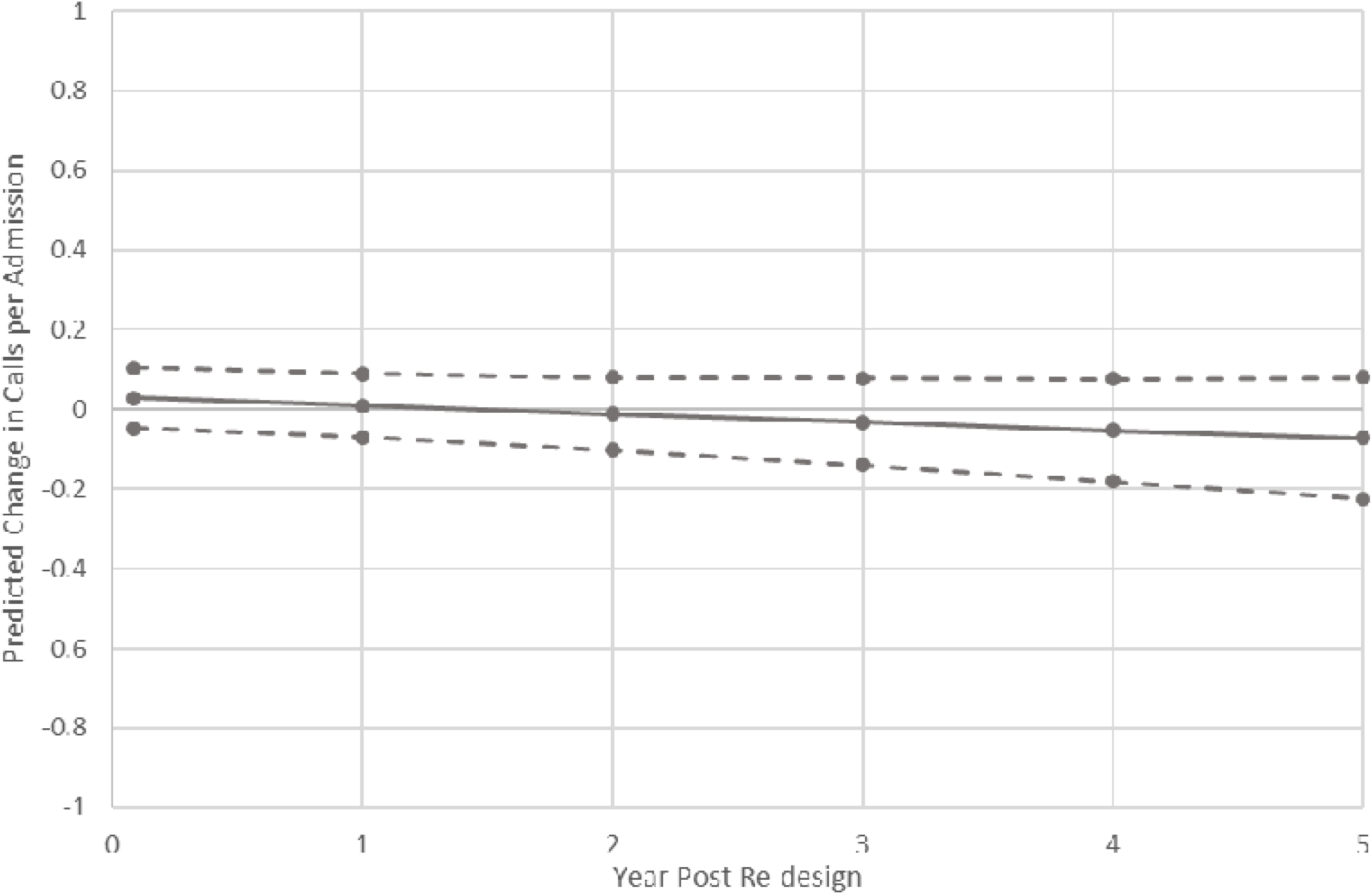
Final Multivariable Model – Cumulative predicted change in calls per admission associated with the RRS re-design, with 95% confidence intervals.

The fit statistics from the ITS models for the calls per admission were similar to those observed in the analysis of the primary outcomes, again suggesting reasonable fit. Durbin’s alternative test statistics were 0.68 on 1 df (P=0.41) and 0.58 on 1 df (P=0.45) for the univariate and multivariable models, respectively. The stationary R^2^ values were 0.26 for the univariate and 0.30 for the multivariable models. The Ljung-Box Q statistic indicated there was no significant lack of fit for the univariate (12.83 on 17 df; P=0.75), nor for the multivariable model (Q=15.82 on 17 df; P=0.54).

A sensitivity analysis excluding the extreme outlier led to results that were essentially unchanged, with the pre-post regression coefficient change in slope of -0.001 [SE 0.001] and 0.07 fewer predicted calls per admission (95%CI -0.21 – 0.07, P=0.34) as seen in the final multivariable model shown in Supplementary Material 2.

## DISCUSSION

### Key Findings

By the end this study, a multi-faceted RRS re-design was associated with a six percent reduction in the percentage of RRT-attended patients having repeat calls and a slight reduction (of 0.07) in the predicted number of RRT calls per patient (having at least one call), versus that predicted, had the re-design not occurred.

Although not statistically significant, these differences in outcome, versus repeat calls predicted in the absence of the re-design, are of potential interest for RRS managers in mitigating potentially preventable RRT calls and planning towards organisational resilience.

### Interpretation of Results

The estimated reduction in proportion of patients experiencing a repeat call following the RRS re-design has potential implications for patient mortality. A previous publication from this project detected a significant association between potentially avoidable repeat calls and in-hospital mortality. [12] Other studies have corroborated the association between repeat calling and mortality more broadly. [30,31] Therefore, while not all repeat calls can be prevented, there is still a strong argument for seeking to minimise the need for their occurrence.

For organisations, there are potential benefits to operational efficiency from reducing the frequency of repeat calling. RRTs tend to draw resources from other acute clinical unit rosters, such as ICU and Internal Medicine, rather than have their own independent staffing. [8,9] Therefore, reductions in potentially avoidable re-calling of the RRT allow staff to attend to their primary rostered clinical duties.

The results in this paper may have practical appeal to RRS managers, as the investigating hospital benefitted by:

- six percent fewer RRT attended patients going on to have repeat calls (per month) by the end of Phase 2 of this study. In the context of a median 30-minute call duration, [12] this equates to a reduction in RRS activity of some three hours per 100 patients attended by the RRT, and
- one fewer call per 14 admissions subject to RRT calling. In the final study year, this would also extrapolate to the hospital reducing potentially preventable RRS activity by 3.5 hours per 100 patients attended.

In hospitals with high call activity, this offsetting of demand may lead to pragmatic gains in RRT availability and, hence, lower the likelihood of requirements to roster multiple concurrent RRTs.

### Contribution to Evidence Base

The literature on re-designing the RRS to improve use of NTS during RRT calls is scant. Most published articles reinforce simulation training as the gold standard mechanism to achieve this. [14,15] Staff training is labour and cost intensive, so alternative strategies need to be explored.

Kansal et al. evaluated streamlining information sharing by ward staff to the RRT on their arrival to calls, alongside other restructuring of their respective RRS. [19] Although they did report reductions in rates of unexpected deaths and other adverse patient outcomes after re-designing the RRS, these authors could not ascribe the role of the enhanced handover as the sole reason for these improvements.

Prince et al. and Mardegan et al. described changes to operations of the RRT during calls. [20,21] Prince et al. focused on visual identification of team member roles during cardiac arrest calls which was incorporated into simulation training for the RRT. These authors noted perceived improvements in communication during RRT calls, although no pre-training data were collected. Mardegan et al. only described staff satisfaction after introduction of a RRT call checklist that facilitated handover at the end of calls. While staff were considered positive in general about each of these interventions, no effects on patient outcomes were presented in either paper. [20,21]

The present study demonstrated feasibility of a multi-faceted RRS re-design to promote use of Non-Technical Skills during RRT calls without the need for costly training. It also presents objective evidence that reductions in repeat RRT calling can be achieved. While not statistically significant, the results are still highly relevant at an organisational level, especially given the negligible barriers or overheads to implementing the three components of this RRS re-design.

### Strengths and Limitations

To the best of the authors’ knowledge, this is the first description of objective RRS performance outcomes arising from implementation of a Non-Technical Skills focused system re-design.

Further, the use of the Interrupted Time Series approach permitted isolating the effect of the re-design on the outcomes. One strength of the analysis is that it accounted for temporal trends and variations. The time-series analysis also suggested that the effect of the re-design in reducing rates of repeat calling was sustained throughout Phase 2, with no evidence of attrition of benefit.

As with any pragmatic study, there are limitations. First, the authors acknowledge the absence of results regarding RRS compliance with the components of the re-design. Due to limited financial resources for the study, it was not possible to employ observers to objectively record attendance at RRT meetings, wearing of badges during RRT calls and adherence to the required hand-off process at RRT call completions. The possibility of having RRT staff collect these data was considered but not used due to wishing to avoid creating additional workload for them at RRT calls, and the potential for missed, incomplete or unreliable datapoints.

Second, although demographic and hospital activity co-variates were included in the analyses and the configuration of RRS did not otherwise alter during the entire study period, it is still possible that some other unmeasured factors were responsible for the findings.

Finally, some repeat calls may indicate a correctly functioning RRS responding to clinically discrete deteriorations. However, this study focused on the wider implication for organisations, and so did not separate these from the preventable calls as all repeat calls present a potential logistical and resourcing burden on hospitals.

### Future Scope for Re-designing the RRS

Using role stickers, rather than badges, and electronic availability of RRT rosters have already been discussed in previous findings from this project. [13]

Further to those, a natural addition to the RRS would be debriefs for the RRT and other hospital staff involved in calls. [33] This could take one of two forms: “hot debrief” conducted immediately after completion of each RRT call or “cold debrief” in which cases are reviewed later at scheduled meetings. [34] Both have arguments in favour and against, but the main barrier is logistical. Hot debrief depends on RRT members, and possibly also ward staff, involved in that call remaining available to attend. For ad-hoc RRTs rostered from other clinical roles, this may be infeasible. [8,9] The scheduled, delayed nature of cold debrief provides more opportunity for RRT members to plan their attendance and avoid conflicts with other clinical duties, so may be easier to implement. [34]

## CONCLUSIONS

This study of a multi-faceted RRS re-design demonstrates its feasibility and a reduction in percentage of patients per month having repeat calls which, while not statistically significant, may be of interest to RRS clinicians and managers.

The findings are indicative of the potential gains from applying cost-effective re-design methodology to address the needs of the RRS.

## Data Availability

The study dataset and the statistical analysis plans are available from the corresponding author on reasonable request. Under the conditions of ethical approval, the study dataset may not be made available publicly.

## ACKNOWLEDGEMENTS

Dr Bill Wilson, Chief Medical Information Officer, Northern Adelaide Local Health Network, Adelaide, Australia for assistance with data extraction from hospital electronic databases

## ETHICS APPROVAL AND PATIENT CONSENT

This study was approved by the Central Adelaide Health Network Human Research Committee, Level 3, Roma Mitchell House, 136 North Terrace, Adelaide, SA 5000, Australia (approval number: 2012069).

The need for patient signed consent was waived on the grounds that data used in this study were already collected electronically for hospital quality assurance purposes, no direct patient identifiers were included in the study database, and all individual patient level variables were aggregated prior to analysis and reporting.

