## Supplement 1 for "Effect of a multi-faceted Rapid Response System re-design on repeat calling of the Rapid Response Team"

Percentage

| Study Month | Pre-Int RepCallAdm |
| --- | --- |
| 1 | 15.3846 |
| 2 | 12.8205 |
| 3 | 16.2162 |
| 4 | 15.5172 |
| 5 | 18.4211 |
| 6 | 20.4082 |
| 7 | 28.2609 |
| 8 | 15.7895 |
| 9 | 15.2174 |
| 10 | 15.625 |
| 11 | 13.8889 |
| 12 | 8.16327 |
| 13 | 17.7778 |
| 14 | 15.6863 |
| 15 | 13.5593 |
| 16 | 5.12821 |
| 17 | 6.66667 |
| 18 | 16.6667 |
| 19 | 17.1429 |
| 20 | 17.2414 |
| 21 | 12.8205 |
| 22 | 22.2222 |
| 23 | 10.3448 |
| 24 | 8.69565 |
| 25 | 13.2075 |
| 26 | 10.5263 |
| 27 | 15.625 |
| 28 | 23.0769 |
| 29 | 16.2791 |
| 30 | 8.33333 |
| 31 | 13.0435 |
| 32 | 16.3636 |
| 33 | 12.5 |
| 34 | 7.5 |
| 35 | 18.5185 |
| 36 | 17.7419 |
| 37 | 26.8657 |
| 38 | 31.4815 |
| 39 | 22.2222 |
| 40 | 28.0702 |
| 41 | 14.8148 |
| 42 | 16.6667 |
| 43 | 12.7273 |
| 44 | 20.4082 |
| 45 | 25.9259 |
| 46 | 19.6429 |
| 47 | 20.339 |
| 48 | 24.3243 |
| 49 | 26.7442 |
| 50 | 24.5902 |
| 51 | 15.942 |
| 52 | 7.69231 |
| 53 | 19.2982 |
| 54 | 19.0476 |
| 55 | 25 |
| 56 | 28.5714 |
| 57 | 18.1818 |
| 58 | 14.2857 |
| 59 | 25.7576 |
| 60 | 15 |

| Study Month | Pre-Int RepCallAdm2 |
| --- | --- |
| 1 | 15.3846 |
| 2 | 12.8205 |
| 3 | 16.2162 |
| 4 | 15.5172 |
| 5 | 18.4211 |
| 6 | 20.4082 |
| 7 | 28.2609 |
| 8 | 15.7895 |
| 9 | 15.2174 |
| 10 | 15.625 |
| 11 | 13.8889 |
| 12 | 8.16327 |
| 13 | 17.7778 |
| 14 | 15.6863 |
| 15 | 13.5593 |
| 16 | 5.12821 |
| 17 | 6.66667 |
| 18 | 16.6667 |
| 19 | 17.1429 |
| 20 | 17.2414 |
| 21 | 12.8205 |
| 22 | 22.2222 |
| 23 | 10.3448 |
| 24 | 8.69565 |
| 25 | 13.2075 |
| 26 | 10.5263 |
| 27 | 15.625 |
| 28 | 23.0769 |
| 29 | 16.2791 |
| 30 | 8.33333 |
| 31 | 13.0435 |
| 32 | 16.3636 |
| 33 | 12.5 |
| 34 | 7.5 |
| 35 | 18.5185 |
| 36 | 17.7419 |
| 37 | 26.8657 |
| 38 | 31.4815 |
| 39 | 22.2222 |
| 40 | 28.0702 |
| 41 | 14.8148 |
| 42 | 16.6667 |
| 43 | 12.7273 |
| 44 | 20.4082 |
| 45 | 25.9259 |
| 46 | 19.6429 |
| 47 | 20.339 |
| 48 | 24.3243 |
| 49 | 26.7442 |
| 50 | 24.5902 |
| 51 | 15.942 |
| 52 | 7.69231 |
| 53 | 19.2982 |
| 54 | 19.0476 |
| 55 | 25 |
| 56 | 28.5714 |
| 57 | 18.1818 |
| 58 | 14.2857 |
| 59 | 25.7576 |
| 60 | 15 |

| Study Month | Post-Int RepCallAdm |
| --- | --- |
| 61 | 23.8095 |
| 62 | 14.1176 |
| 63 | 16.25 |
| 64 | 22 |
| 65 | 26.3158 |
| 66 | 18.4615 |
| 67 | 18.6441 |
| 68 | 25.7576 |
| 69 | 13.9535 |
| 70 | 23.7288 |
| 71 | 25.641 |
| 72 | 36.7647 |
| 73 | 22.7273 |
| 74 | 15.8537 |
| 75 | 28.4091 |
| 76 | 21.1765 |
| 78 | 21.7949 |
| 79 | 20.6186 |
| 80 | 18.8889 |
| 81 | 22.7273 |
| 82 | 30 |
| 83 | 20.9091 |
| 84 | 19.685 |
| 85 | 17.2414 |
| 86 | 19.1304 |
| 87 | 20.5357 |
| 88 | 18.75 |
| 89 | 18.018 |
| 90 | 22.549 |
| 91 | 23.7624 |
| 92 | 23.7624 |
| 93 | 12.7273 |
| 94 | 25 |
| 95 | 15.3061 |
| 96 | 25.5814 |
| 97 | 15 |
| 98 | 24.375 |
| 99 | 22.293 |
| 100 | 28.1818 |
| 101 | 25.3731 |
| 102 | 29.5918 |
| 103 | 20.7921 |
| 104 | 20.6897 |
| 105 | 25.3846 |
| 106 | 24.8 |
| 107 | 23.1788 |
| 108 | 26.6187 |
| 109 | 22.6027 |
| 110 | 24.8276 |
| 111 | 21.831 |
| 112 | 19.0476 |
| 113 | 29.6296 |
| 114 | 20.3252 |
| 115 | 23.2558 |
| 116 | 22.6891 |
| 117 | 22.5 |
| 118 | 23.1884 |
| 119 | 22.5564 |
| 120 | 27.6316 |

Percentage of repeat call admissions per month representing the ARIMA univariate model with outlier detection enabled. Phase 1 observed data shown as black squares, with trend shown as the solid black line. Phase 2 observed data shown as grey circles, with trend shown as the solid grey line. The Phase 1 trend is extended into Phase 2 as the dotted black line for comparison with Phase 2 observed data. Observation for study month 77 (November 2015) was identified as an outlier and excluded for this sensitivity analysis.
