## Supplement 2 for "Effect of a multi-faceted Rapid Response System re-design on repeat calling of the Rapid Response Team"

Mean

| Study Month | Pre-Int CallPerAdm |
| --- | --- |
| 1 | 1.17949 |
| 2 | 1.12821 |
| 3 | 1.16216 |
| 4 | 1.36207 |
| 5 | 1.26316 |
| 6 | 1.26531 |
| 7 | 1.43478 |
| 8 | 1.18421 |
| 9 | 1.15217 |
| 10 | 1.1875 |
| 11 | 1.13889 |
| 12 | 1.08163 |
| 13 | 1.22222 |
| 14 | 1.23529 |
| 15 | 1.28814 |
| 16 | 1.05128 |
| 17 | 1.06667 |
| 18 | 1.2 |
| 19 | 1.17143 |
| 20 | 1.24138 |
| 21 | 1.17949 |
| 22 | 1.33333 |
| 23 | 1.12069 |
| 24 | 1.1087 |
| 25 | 1.18868 |
| 26 | 1.21053 |
| 27 | 1.21875 |
| 28 | 1.28205 |
| 29 | 1.18605 |
| 30 | 1.11111 |
| 31 | 1.15217 |
| 32 | 1.18182 |
| 33 | 1.125 |
| 34 | 1.075 |
| 35 | 1.24074 |
| 36 | 1.19355 |
| 37 | 1.46269 |
| 38 | 1.40741 |
| 39 | 1.26667 |
| 40 | 1.35088 |
| 41 | 1.18519 |
| 42 | 1.16667 |
| 43 | 1.16364 |
| 44 | 1.2449 |
| 45 | 1.2963 |
| 46 | 1.28571 |
| 47 | 1.40678 |
| 48 | 1.31081 |
| 49 | 1.40698 |
| 50 | 1.32787 |
| 51 | 1.26087 |
| 52 | 1.07692 |
| 53 | 1.26316 |
| 54 | 1.2381 |
| 55 | 1.39286 |
| 56 | 1.42857 |
| 57 | 1.25758 |
| 58 | 1.17857 |
| 59 | 1.37879 |
| 60 | 1.225 |

| Study Month | Pre-Int CallPerAdm2 |
| --- | --- |
| 1 | 1.17949 |
| 2 | 1.12821 |
| 3 | 1.16216 |
| 4 | 1.36207 |
| 5 | 1.26316 |
| 6 | 1.26531 |
| 7 | 1.43478 |
| 8 | 1.18421 |
| 9 | 1.15217 |
| 10 | 1.1875 |
| 11 | 1.13889 |
| 12 | 1.08163 |
| 13 | 1.22222 |
| 14 | 1.23529 |
| 15 | 1.28814 |
| 16 | 1.05128 |
| 17 | 1.06667 |
| 18 | 1.2 |
| 19 | 1.17143 |
| 20 | 1.24138 |
| 21 | 1.17949 |
| 22 | 1.33333 |
| 23 | 1.12069 |
| 24 | 1.1087 |
| 25 | 1.18868 |
| 26 | 1.21053 |
| 27 | 1.21875 |
| 28 | 1.28205 |
| 29 | 1.18605 |
| 30 | 1.11111 |
| 31 | 1.15217 |
| 32 | 1.18182 |
| 33 | 1.125 |
| 34 | 1.075 |
| 35 | 1.24074 |
| 36 | 1.19355 |
| 37 | 1.46269 |
| 38 | 1.40741 |
| 39 | 1.26667 |
| 40 | 1.35088 |
| 41 | 1.18519 |
| 42 | 1.16667 |
| 43 | 1.16364 |
| 44 | 1.2449 |
| 45 | 1.2963 |
| 46 | 1.28571 |
| 47 | 1.40678 |
| 48 | 1.31081 |
| 49 | 1.40698 |
| 50 | 1.32787 |
| 51 | 1.26087 |
| 52 | 1.07692 |
| 53 | 1.26316 |
| 54 | 1.2381 |
| 55 | 1.39286 |
| 56 | 1.42857 |
| 57 | 1.25758 |
| 58 | 1.17857 |
| 59 | 1.37879 |
| 60 | 1.225 |

| Study Month | Post-Int CallPerAdm |
| --- | --- |
| 61 | 1.32143 |
| 62 | 1.2 |
| 63 | 1.2875 |
| 64 | 1.3 |
| 65 | 1.26316 |
| 66 | 1.2 |
| 67 | 1.22034 |
| 68 | 1.39394 |
| 69 | 1.30233 |
| 70 | 1.30508 |
| 71 | 1.32051 |
| 72 | 1.47059 |
| 73 | 1.33636 |
| 74 | 1.17073 |
| 75 | 1.44318 |
| 76 | 1.25882 |
| 78 | 1.29487 |
| 79 | 1.39175 |
| 80 | 1.24444 |
| 81 | 1.30909 |
| 82 | 1.45 |
| 83 | 1.5 |
| 84 | 1.29921 |
| 85 | 1.31034 |
| 86 | 1.26087 |
| 87 | 1.27679 |
| 88 | 1.28125 |
| 89 | 1.25225 |
| 90 | 1.29412 |
| 91 | 1.30693 |
| 92 | 1.35644 |
| 93 | 1.22727 |
| 94 | 1.29167 |
| 95 | 1.23469 |
| 96 | 1.37984 |
| 97 | 1.19167 |
| 98 | 1.40625 |
| 99 | 1.35669 |
| 100 | 1.35455 |
| 101 | 1.3806 |
| 102 | 1.44898 |
| 103 | 1.33663 |
| 104 | 1.30172 |
| 105 | 1.35385 |
| 106 | 1.328 |
| 107 | 1.39073 |
| 108 | 1.42446 |
| 109 | 1.30822 |
| 110 | 1.42759 |
| 111 | 1.33099 |
| 112 | 1.2449 |
| 113 | 1.41481 |
| 114 | 1.28455 |
| 115 | 1.34109 |
| 116 | 1.40336 |
| 117 | 1.29167 |
| 118 | 1.30435 |
| 119 | 1.28571 |
| 120 | 1.48026 |

Mean number of calls per admission by study month for the ARIMA univariate model with outlier detection enabled. Phase 1 observed data shown as black squares, with trend shown as the solid black line. Phase 2 observed data shown as in grey circles, with trend shown as the solid grey line. The Phase 1 trend is extended into Phase 2 as the dotted black line for comparison with Phase 2 observed data. Observation for study month 77 (November 2015) was identified as outlier and excluded for this sensitivity analysis.
